# Implementation of Early Intervention in Psychosis Initiatives in Latin America and the Caribbean: A Case Study

**DOI:** 10.1101/2025.11.11.25340030

**Authors:** Ruben Valle, Camila Velez, Srividya N. Iyer

**Author notes:** **Corresponding author:** (SNI), ruben, (RV).

## Abstract

Psychosis is a serious mental illness, with onset in adolescence and young adulthood. Few early intervention in psychosis (EIP) programs exist in the Global South, where most of the world’s youth live. Addressing this gap requires understanding implementation contexts, pathways and challenges. This study examines EIP initiatives in Latin America and the Caribbean (LAC) and explores implementers’ perspectives on scaling them. A single-case study design was employed. Guided by the Exploration, Preparation, Implementation, and Sustainment (EPIS) framework, we conducted semi-structured interviews with EIP implementers across LAC and gathered policy documents. Data was coded and analyzed using thematic analysis. Twenty-five participants from 10 countries described 26 initiatives, including clinical and research programs, guidelines, and a technical standard. Themes were mapped onto the EPIS framework. In Exploration, participants highlighted key motivators, the influence of collaborations with foreign researchers, contextual adversity (e.g., poverty, stigma), and the role of Indigenous cosmologies and religious traditions in shaping care pathways. In Preparation, they emphasized difficulties in culturally adapting models from high-income countries (HICs), limited staff awareness, and resource shortages. In Implementation, participants described how initiatives operated in local contexts (e.g., research programs offering care to address unmet needs), how they were generally well received by patients and staff, and the shortage of psychosocial interventions. In sustainment, few initiatives persisted; participants pointed to dependence on international funding, limited policy support, capacity, and awareness. While EIP was valued, national dissemination of HIC-based programs was considered unfeasible. EIP development in LAC has occurred amid structural and resource limitations affecting many LMICs. Implementers’ proposals: task-shifting; simplified care packages; leveraging extant services; and enhancing early psychosis literacy— represent feasible strategies to support EIP across LAC. Recommendations for future research, including the involvement of service users and their families and the adaptation of implementation frameworks to context, are shared.

## Introduction

Early Intervention in Psychosis (EIP) is a multicomponent service model for early stages of psychotic disorders, specifically, clinical high risk for psychosis (CHR) and first-episode psychosis (FEP) [1,2]. Grounded in a philosophy of hope and optimism [3], EIP promotes recovery by offering targeted interventions to patients and families (i.e., medication, case management, cognitive-behavioral therapy, family psychoeducation, etc.) [2]. Evidence from randomized clinical trials, systematic reviews, and meta-analyses shows that EIP leads to better clinical and functional outcomes in FEP compared to standard care [4–7]. Cost-effectiveness analyses demonstrate benefits in high- and low-resource countries [8,9]. For CHR, evidence is relatively weaker but suggests that EIP can reduce symptoms and potentially delay or prevent onset [10].

EIP programs are widely implemented in many high-income countries (HICs) [11], typically as stand-alone services, but also through integrated services and hub-and-spoke models [12]. Pathways to implementation have been diverse, including government policies and research projects [13]. The success of EIP in HICs has relied on leadership, sustained funding, supportive policies, and partnerships [11]. Efforts complementary to EIP services development, like research [14], guidelines [15], and policies, have helped generate evidence, standardize care, and raise awareness about EIP’s benefits [16].

In Latin America and the Caribbean (LAC), where almost 60% of countries are low-and middle-income countries (LMICs) [17], EIP implementation has been fragmented. Brazil established the first documented EIP program in 1999 [18], contemporaneous with many HICs. Yet by 2011, a narrative review found that only Brazil and Mexico had programs [19]. A 2020 scoping review identified EIP programs in those two countries, plus two Chilean facilities and a one-off Argentinian study training primary care staff to refer FEP cases [20]. A 2025 narrative review reported no further expansion [21]. EIP programs in LAC remain concentrated in research centers within tertiary care in metropolitan areas, thus reaching fewer people [19–21].

Previous work on EIP in LAC has largely used desk reviews and focused on clinical programs [19–21], limiting our understanding into contextual factors, implementation processes, or challenges underlying uneven provision and uptake. Non-clinical initiatives have been overlooked despite their potential role in expanding EIP. For instance, many research projects in LMICs aim to evolve into services [22,23], and policies can facilitate care provision in psychosis [24]. Replicating EIP programs from HICs in LAC is not feasible due to socio-cultural and resource differences, highlighting the need to examine the full spectrum of EIP initiatives and multiple implementation pathways [25,26].

Addressing these gaps requires applying well-established implementation science frameworks to analyze implementation pathways, strategies, and contextual influences, as well as the roles and motivations of diverse stakeholders advancing EIP within their countries. Accordingly, this study’s objectives are to examine the implementation processes of EIP initiatives in LAC and explore implementers’ perspectives on disseminating EIP across the region.

## Methods

### Design

This qualitative study uses a single case design with embedded units [27]. The single case is EIP implementation in LAC. The embedded units of analysis are country-level EIP initiatives. This design was chosen to generate an in-depth analysis of the implementation of EIP in LAC as a region, and each initiative’s trajectory. The study was guided by the well-established [28,29] Exploration, Preparation, Implementation, and Sustainment (EPIS) implementation framework [30–32]. This framework examines outer, inner, bridging, and intervention contexts and different implementation phases, and is therefore well aligned with the study’s objectives. EPIS phases are: *Exploration (*assessing needs and deciding to adopt the intervention), Preparation (identifying barriers, facilitators, and adaptations), *Implementation* (launching the intervention and monitoring) and *Sustainment* (ensuring continued delivery). The study follows the Standards for Reporting Qualitative Research (S1 Table) [33]. The project was approved by McGill University’s Institutional Review Board (reference number A05-E47-24B; approved on May 30, 2024). All participants provided written informed consent and agreed to the audio and/or video recording of interviews. Data collection occurred between August 23, 2024, and February 28, 2025.

### Definitions

An EIP initiative refers to a plan or process addressing FEP or at CHR, including:

- Clinical programs: organized healthcare services providing assessment, treatment, and follow-up.
- Research programs: structured, often multi-year initiatives, to generate knowledge around a theme.
- Clinical guidelines: evidence-based recommendations developed by experts for assessment, diagnosis and treatment.
- Technical standards: documents issued by an authoritative body specifying procedures, minimum requirements, and operational criteria for service delivery.
- Individual studies: standalone research projects addressing specific questions or evaluating care components.

### Study Settings

LAC has 664 million inhabitants in 42 countries [34]; 24.5% are aged 15-29 years [34], an age group at heightened risk for psychosis [35]. By 2015, Brazil, Chile, Panama, and Peru had community-based mental health models [36,37]; in most other countries, care remains centralized in city-based psychiatric hospitals [38]. Schizophrenia affects 277.8 per 100,000 people in LAC [39], yet estimated service coverage is only 26.7% [40]. Treatment consists primarily of antipsychotic medication, with limited availability of psychosocial interventions [41].

### Participants

Eligible participants were clinicians, researchers, or policymakers involved in implementing EIP initiatives in LAC. Purposive sampling [42] was used to identify initial implementers via systematic reviews [19–21], conference abstracts [43], and networking. We compiled a dataset of implementers (initiative type, implementers’/authors’ names, institutions, emails). We contacted participants via email to explain the study, obtain consent and schedule the individual interview. We used snowball sampling to identify additional participants.

### Sample size

The sample size was determined based on data saturation [44] and ensuring the inclusion of at least one implementer per identified initiative. The final sample included 25 participants — 20 of the 22 initially identified implementers (response rate=86.7%), two replacements for non-respondents (e.g., co-authors) and three identified through snowball sampling. Data saturation was reached by interview 22; but three subsequent interviews covered less common initiatives. Table 1 presents sample characteristics.

**Table 1.**
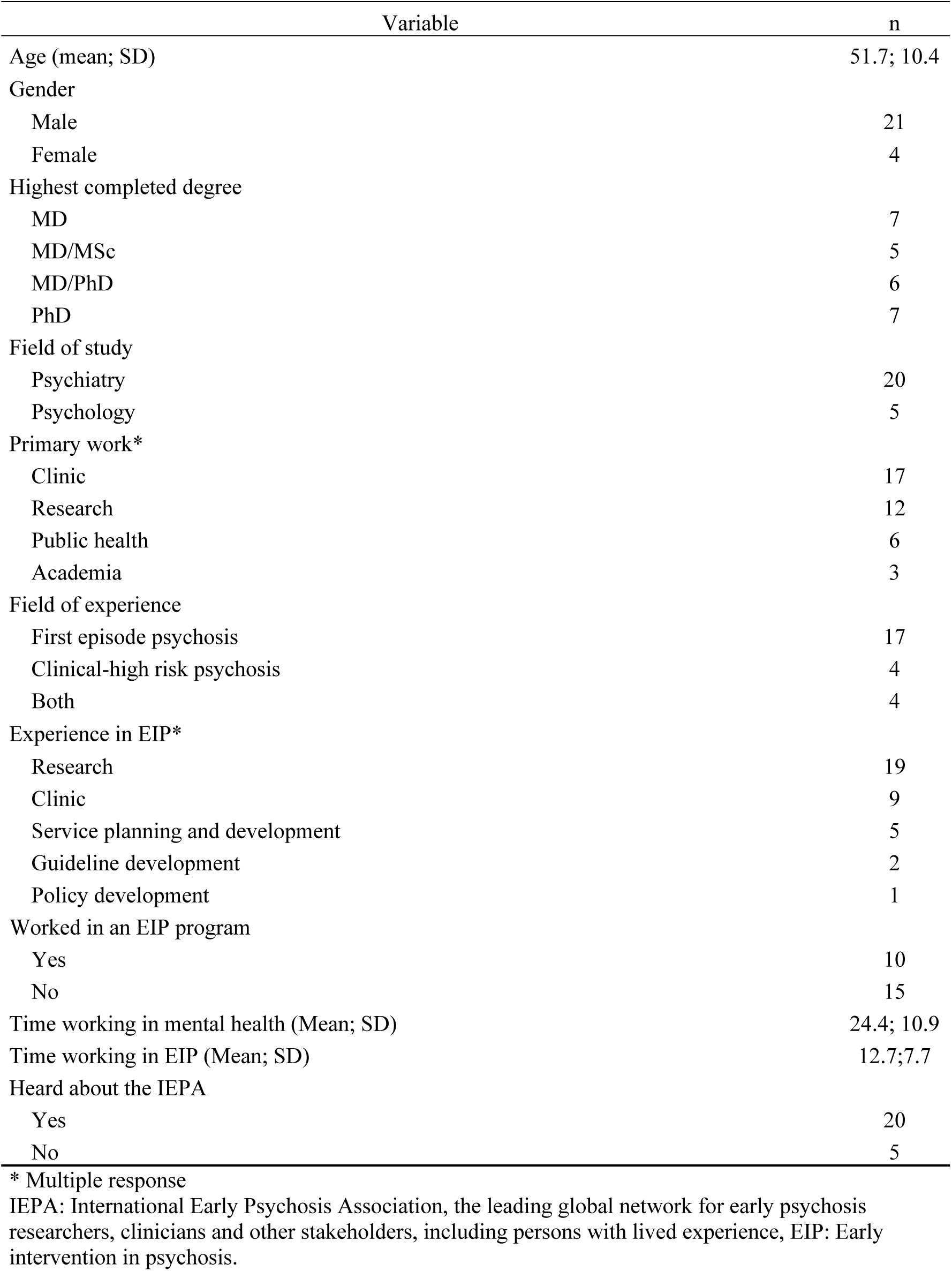
Sociodemographic variables of participants (n=25).

| Variable | n |
| --- | --- |
| Age (mean; SD) | 51.7; 10.4 |
| Gender |  |
| Male | 21 |
| Female | 4 |
| Highest completed degree |  |
| MD | 7 |
| MD/MSc | 5 |
| MD/PhD | 6 |
| PhD | 7 |
| Field of study |  |
| Psychiatry | 20 |
| Psychology | 5 |
| Primary work* |  |
| Clinic | 17 |
| Research | 12 |
| Public health | 6 |
| Academia | 3 |
| Field of experience |  |
| First episode psychosis | 17 |
| Clinical-high risk psychosis | 4 |
| Both | 4 |
| Experience in EIP* |  |
| Research | 19 |
| Clinic | 9 |
| Service planning and development | 5 |
| Guideline development | 2 |
| Policy development | 1 |
| Worked in an EIP program |  |
| Yes | 10 |
| No | 15 |
| Time working in mental health (Mean; SD) | 24.4; 10.9 |
| Time working in EIP (Mean; SD) | 12.7; 7.7 |
| Heard about the IEPA |  |
| Yes | 20 |
| No | 5 |

\* Multiple response
IEPA: International Early Psychosis Association, the leading global network for early psychosis researchers, clinicians and other stakeholders, including persons with lived experience, EIP: Early intervention in psychosis.

Seventeen participants reported research as their primary activity and 12 clinical practice. 20 had backgrounds in psychiatry and five in psychology.

### Data collection

Semi-structured interviews (∼one hour) were conducted via Zoom in Spanish or English and were video-recorded. We used a demographic questionnaire and an EPIS-based open-ended interview guide, that was adapted to each EIP initiative and topics raised during interviews. The questionnaire and guide were piloted in three interviews with EIP coordinators in Canada and India. At the end of the interview, participants were asked to share relevant policy documents.

### Reflexivity

RV, the first author and interviewer, is a male psychiatrist from Peru with a master’s in epidemiology and a doctoral focus on EIP. His professional background and cultural proximity may have facilitated rapport and trust with interviewees. CV, an immigrant woman from Colombia, is a psychotherapist and doctoral student with no prior EIP experience, offering an outsider perspective and approaching the data with fresh eyes. SNI, an immigrant psychologist and experienced EIP researcher in Canada and LMICs, contributed broad expertise during analysis. We remained attentive throughout to how our identities and positionalities influenced interactions, theme development, and interpretation.

### Data analysis

Recordings were transcribed in their original language. Following an iterative process during and after data collection, we used thematic analysis to identify, analyze, and report patterns within the data [45]. This involved familiarization with the data, generation of initial codes, identification and review of themes and subthemes, refinement, definition, and synthesis. RV and CV independently coded transcripts in ATLAS.ti (v25), met regularly and reached consensus through discussion, involving senior author SNI at critical junctures. Themes were subsequently mapped onto the EPIS framework, which are presented in Results with illustrative quotes.

Rigor and trustworthiness [46] were ensured through prolonged engagement with the data, investigator triangulation via independent coding, data triangulation between interviews and four health policy documents shared by participants, grounding findings in participants’ quotations, and member checking by sharing preliminary themes with participants (in group presentationsand individual communications).

## Results

### EIP initiatives

Participants implemented 26 EIP initiatives across 10 countries, with one participant reporting on two (Table 2). These included 11 clinical programs, seven research programs, two clinical guidelines, one technical standard, and five individual studies. Most (19) initiatives focused on FEP, five on CHR, and two on both. All progressed through the exploration and preparation phases of EPIS. At the time of the interviews, one standard awaited government approval, and one planned clinical program awaited funding for implementation. Among those implemented, only some advanced to sustainment. Two clinical programs were discontinued, four studies ended without follow-up projects, two research programs concluded without additional funding, and one study and one research program were ongoing but faced uncertain futures (Fig 1).

**Fig 1.**
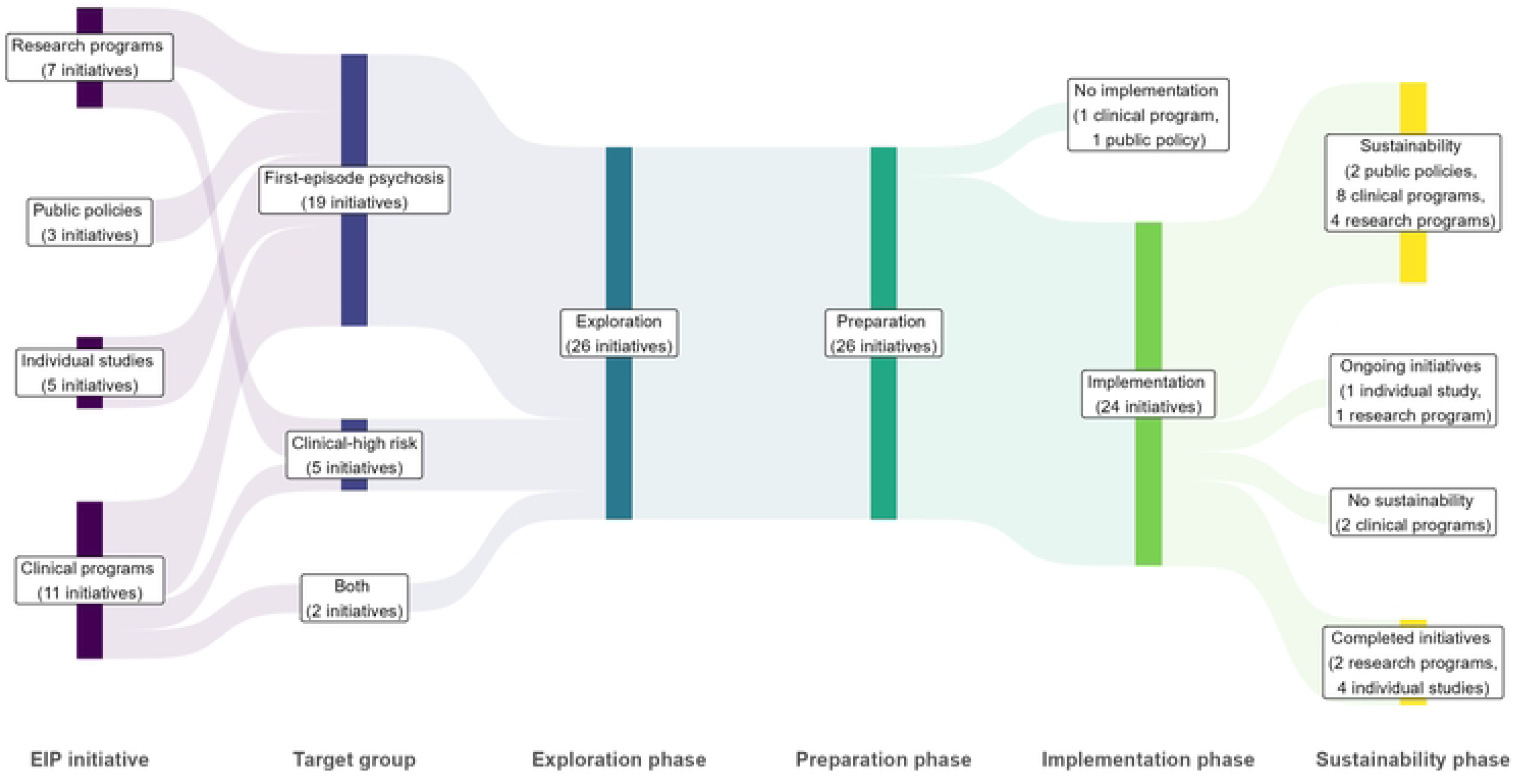
Implementation trajectories of EIP initiatives in Latin America and the Caribbean (n=26 EIP initiatives). Public policies include clinical guidelines and standards.

**Table 2.** Early intervention in psychosis initiatives by country (n=26).

| Country | Income level | “Individual” EIP study | Research program | Clinical program | Clinical guideline | Technical standard | Total |
| --- | --- | --- | --- | --- | --- | --- | --- |
| Country 1 | UMIC |  | 1 initiative | 5 initiatives |  |  | 6 |
| Country 2 | HIC | 1 initiative | 1 initiative | 2 initiatives | 1 initiative |  | 5 |
| Country 3 | UMIC | 1 initiative | 2 initiatives | 2 initiatives |  |  | 5 |
| Country 4 | HIC |  |  | 2 initiatives |  |  | 2 |
| Country 5 | HIC |  | 1 initiative |  |  |  | 1 |
| Country 6 | UMIC | 2 initiatives |  |  |  |  | 2 |
| Country 7 | LMIC |  |  | 1 initiative |  |  | 1 |
| Country 8 | UMIC | 1 initiative |  |  |  |  | 1 |
| Country 9 | UMIC |  |  |  | 1 initiative | 1 initiative | 2 |
| Country 10 | HIC | 1 initiative |  |  |  |  | 1 |

LMIC: Lower-middle income country, UMIC: Upper-middle income country, HIC: High-income country. EIP: Early intervention in psychosis.

### Exploration phase

Participants reflected on their motivations for implementing EIP and their local settings (Fig 2). Most initiatives were locally driven, motivated by participants’ awareness of the impacts of untreated psychotic illnesses and unmet needs for comprehensive care on individuals and their families (suffering, employment/social losses, etc.). Many became involved in EIP through attending international meetings, prior work in non-EIP psychosis services, and/or postgraduate or international training.

> *“The World Psychiatric Association offered training opportunities. I applied for a call to receive training at [organization in HIC], where they kindly showed me everything they were doing and allowed me to participate in their activities. This greatly inspired me to propose an early intervention program.” [Interview 5]*

**Fig 2.**
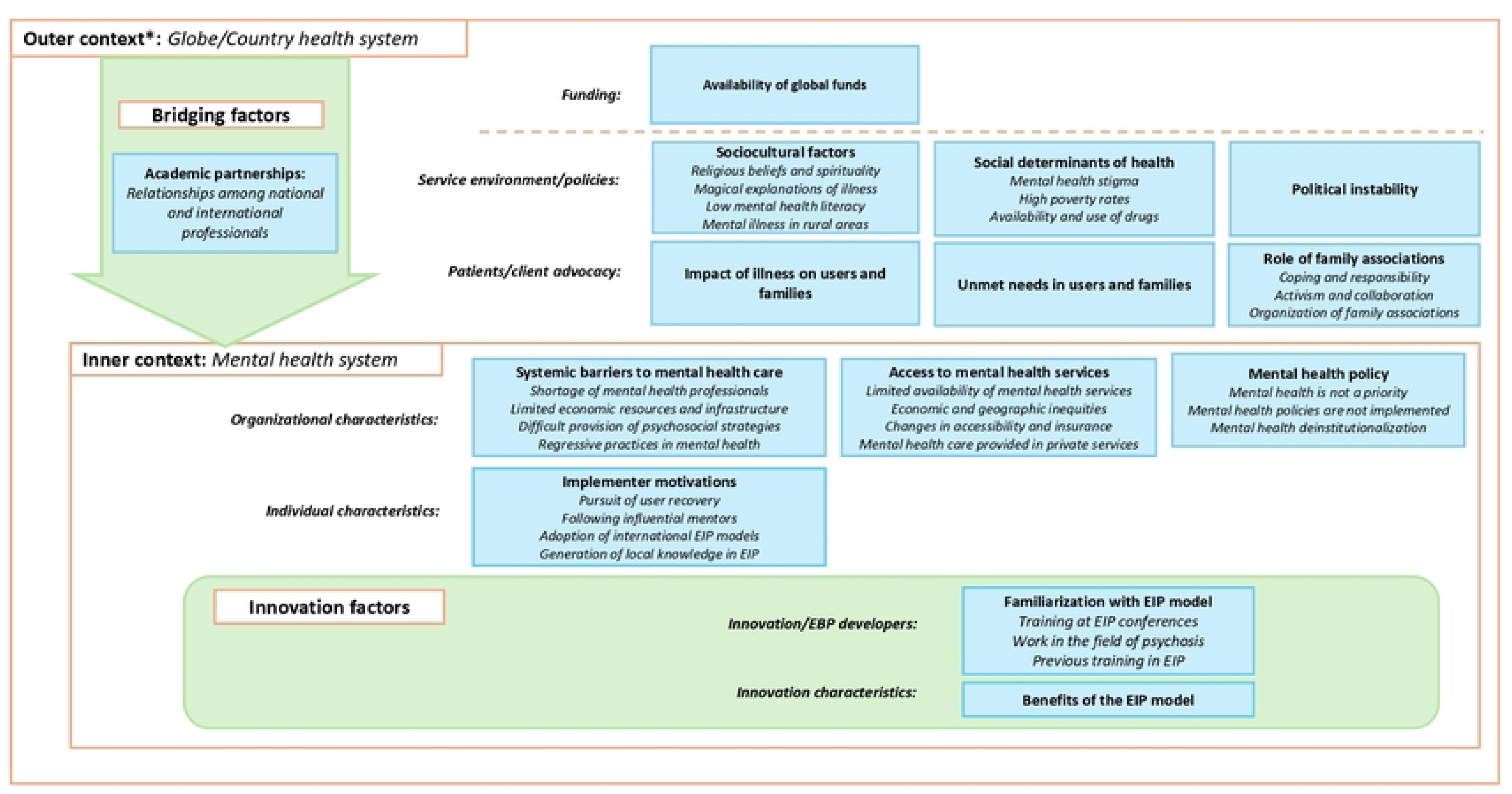
Themes and subthemes identified during the Exploration phase of EIP initiatives (n = 26). *The outer context comprises two dimensions, separated by the dashed orange line: global context and national health system.

Clinicians were inspired by EIP’s recovery orientation and by mentors. Researchers were drawn to generating local evidence for EIP in LAC, while policymakers aimed to implement best practices for people with psychosis. In a few cases, international researchers and funders initiated projects in collaboration with local leads, with funders requiring locally based leadership.

> *“Our program, in contrast, emerged because a group at [foreign university], led by [researcher], a professor of psychiatric epidemiology with experience working in LAC, reached out to us and said it might be interesting to implement an adapted version of [EIP program] in [country], taking advantage of the [Health policy] to develop a program specifically for first-episode psychosis. So, we applied for an [foreign agency] grant. In that context, we adapted and evaluated [EIP program].” [Interview 2]*

Implementation contexts were widely seen as challenging. Mental health was not a governmental priority; recent regulations, though well-intentioned, often failed due to limited resources and organizational capacity; and deinstitutionalization policies were rarely paired with services development. Only one country, where a clinical guideline for first-episode schizophrenia is supported by a universal access to health care law and strong primary care, was seen as supportive of case identification. Most described persistent structural barriers, including shortages of personnel, infrastructure, funding, and capacity for psychosocial care and outdated practices such as pro-institutionalization policies in some areas. Despite reforms and expanded insurance coverage, access remained limited, unequal, and often dependent on private services.

> *“… what happens in [country], especially in cities without academic services or emergency psychiatric units, is that people with FEP experience a longer duration of untreated psychosis, because they don’t have easy access to these facilities.” [Interview 8]*

Participants also identified sociocultural factors which shaped local understandings of mental illness and help-seeking and care pathways, such as low mental health literacy, stigma, supernatural beliefs, like *Aluxes (supernatural beings in Maya cosmology)*, and strong religious traditions such as Catholicism, Kardecist spiritism and African religions. Individuals or families often consulted shamans, priests, or healers before formal services.

> *“So many religions in [country] involve spiritual contact with dead people, so it’s not always easy to tell whether a patient’s symptoms are due to psychosis or are part of their cultural background. This can interfere people from seeking psychiatric help, as they may first consult a priest or a medium associated with these religions.” [Interview 17]*

Many participants identified adverse social determinants disproportionately affecting youths in LAC, including poverty, violence, and the availability of drugs. Political instability in some countries further disrupted health policy coordination. Within these challenging contexts, families played central, complex roles, sometimes seeking institutionalization due to pessimism about recovery, but more often acting as recovery allies and advocates working with government and health institutions to support individuals with mental health conditions. In rural areas, family support, being in nature and working in the field (e.g., herding sheep) were identified as recovery-promoting.

> *“I tell you an anecdote about a young man who, before coming to our service, visited another psychiatric service. He was in his final years of secondary school when he started to suffer from psychosis. The doctor’s message to the mother was: ‘Madam, take him out of school, don’t waste your money and your time, this boy, with difficulty in 10 years will be able to say his name’. She turned out to be a very brave mother and continued to seek treatment. Not only did she encourage him to continue in school, but she also encouraged the young man to enroll in university…This young man applied to law school, beating out a lot of people of his generation.” [Interview 5]*

### Preparation phase

In the preparation phase, local actors developed EIP initiatives in clinical services, universities, and/or public agencies, these settings shaping their focus and structure (Fig 3). Participants affiliated with health institutions created EIP programs that integrated clinical and research components; public agencies concentrated on developing policies; and university researchers led studies or contributed to guidelines. Except for public agencies developing policies, most relied on individual initiative and strong leadership, using strategies like engaging decision-makers and building institutional relationships.

**Fig 3.**
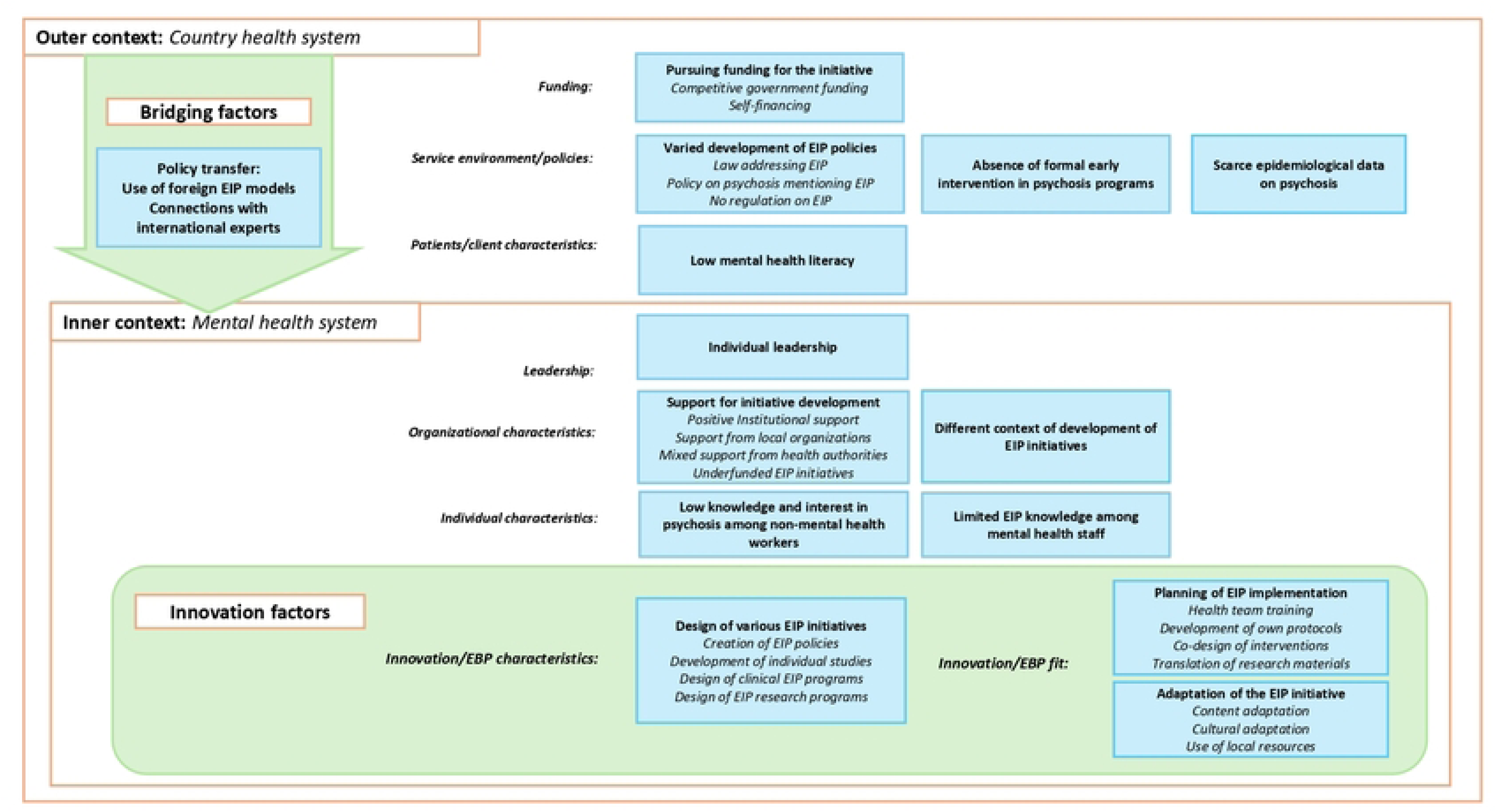
Themes and subthemes identified during the Preparation phase of EIP initiatives (n = 26).

> *“They told me that there were psychiatrists who might be interested in this subject and then they passed me the details of the director of the schizophrenia clinic, and I made an appointment with him and his team…They helped me a lot to open doors… So, you meet one person, he gets involved in the project, and then you get to know someone else.” [Interview 1]*

Another key step was adapting EIP initiatives, as most were based on models from HICs, e.g., Australia or U.S.A., reflecting participants’ training, work experience, or institutional ties. A common strategy was to preserve core evidence-based components, use local resources, and align with public health priorities. For instance, one participant emphasized strengthening family interventions, as youths in their context often live with their parents well into adulthood, reflecting the value of *familismo* (family unity, obligation, and interconnection) common in LAC. Cultural adaptation was often considered but applied unevenly due to its complexity. Common adaptations were translating tools, involving traditional healers, and incorporating cultural activities.

> *“I don’t think people have a good definition of cultural adaptation. There are models, I know several, FRAME is one of them. ADAPT is another one that always includes that cultural element. But the cultural, I think is difficult, how to operate it…[Interview 2]*

Participants described institutional support as generally positive, though seldom accompanied by additional resources. Several initiatives received backing from professional associations and universities. Some required collaboration with health authorities, usually the Ministry of Health, which was mostly a positive process, with some exceptions. Funding sources varied: clinical and government programs often used regular resources, while research-oriented initiatives relied on competitive grants. This was seen as challenging, requiring repeated applications to sustain activities.

> *“The initial program was based on research funds, with competitive research funds here in [country]. Until this year, science was done based on these competitive funds. From one of those research funds, this program was put together…because the clinic lent us the facilities, but the human resource to be able to evaluate and follow up these patients was what the funds mainly financed”. [Interview 9]*

External factors also shaped the planning and design of EIP initiatives. In one country, EIP was backed by a clinical guideline for first-episode schizophrenia, and in a few others, clinical guidelines for psychosis included EIP, but most lacked formal policies. Participants noted the absence of dedicated services for early psychosis, making these initiatives pioneering efforts. Designing and integrating these programs was difficult due to scarce epidemiological data, low mental health literacy and limited capacity or interest among non-mental health professionals. Several noted that EIP was a new concept even for most mental health staff.

> *“When the training was done, it was seen that this [EIP] was something relatively new, even for psychiatrists… I understand, at least when I graduated and was an undergraduate, I finished in 2018, I remember that there was no talk of a first psychotic episode as such until that date.” [Interview 3]*

Preparatory actions included staff training, translating international research materials, and developing care protocols from HICs’ manuals. Policy initiatives often co-designed documents with user and family associations. Two initiatives had not been implemented at the time of interviews due to funding challenges, highlighting barriers to implementation.

> *“Yes, we presented it [EIP clinical program] to the hospital, presented it to the Ministry of Health, then to the [country] Society of Psychiatry. We even applied for a Canadian fund, but we didn’t manage to get funding. We applied to two international calls for proposals and were unable to move forward due to lack of financial resources.” [Interview 5]*

### Implementation phase

Participants described how EIP initiatives functioned in their settings, were received by users and providers, and the challenges faced (Fig 4). In the country with the clinical guideline for first-episode schizophrenia and strong primary care, the policy provided a strong foundation for further initiatives. Individual studies addressed specific goals but did not expand into broader research agendas. Clinical programs focused on care delivery, though they also engaged in research. Research programs initially prioritized scientific inquiry, but those in clinical settings often began offering care to address unmet needs. As a result, most programs combined care and research to varying extents.

**Fig 4.**
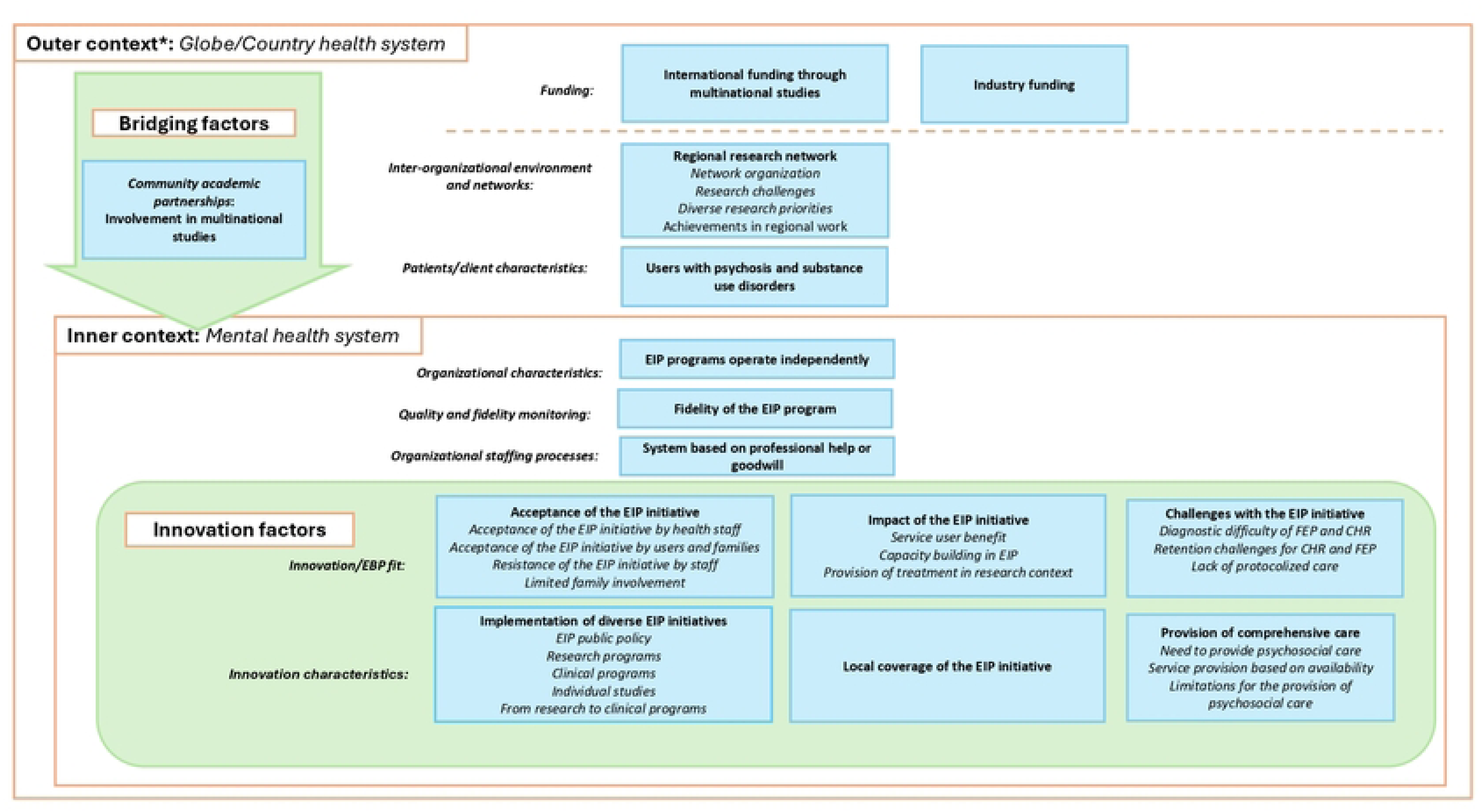
Themes and subthemes identified during the Implementation phase of EIP initiatives (n = 26). *The outer context comprises two dimensions, separated by the dashed orange line: global context and national health system.

“*We first initiated as a research program, so at the beginning we only had the assessments and the medical and then after the research assessments…[we began] to provide care, but we were all psychiatrists. So, I would say that things were evolving as patients were being enrolled in the research and we needed to provide some care for them.*” *[Interview 8]*

Participants described their clinical and research EIP initiatives as well-received by users and families, who trusted implementers’ clinical competence or institutional reputation. However, initial enthusiasm sometimes declined, due to limited family involvement, unrealistic expectations for rapid recovery or rising comorbidity with substance use, which demanded more complex clinical management. While EIP models were generally accepted by mental health providers in these contexts, some resistance to innovation and the perception that there is no difference between FEP and chronic stages were reported. Despite challenges, initiatives were generally seen as clinically important and key to capacity-building. Research programs also enabled early detection and treatment of cases that might otherwise go missed.

> *“In our research, sometimes we did anti-psychotics for them. Sometimes we did antidepressants, sometimes we referred them to the psychologist for psychotherapy. Sometimes this was done for individual psychotherapy group therapy, and sometimes we just maintained surveillance on symptoms. So this was not structured, but was more on a as needed basis on an individual basis.” [Interview 6]*

Across all initiatives, psychosocial interventions were deemed important but difficult to sustain due to resource and trained staffing shortages. Only one research program on psychosis epidemiology prioritized case identification over treatment but noted some psychosocial interventions offered within local services. Psychosocial interventions specialized for psychosis, including cognitive-behavioral therapy, were rarely available. Reflecting on constraints, one participant noted they could deliver only half of what might be clinically recommended for psychosis.

> *“I think it [referring to fidelity scale for EIP] was developed by Donald Addington, and we realized half of what would be needed to be a first episode of psychosis proper program. But this is due to the lack of support…I think that we are not much better than we were at the beginning.” [Interview 8]*

Participants described clinical and research initiatives as often disconnected from the broader health system. Private sector programs served small groups; public programs also had low coverage due to their stand-alone structure. In contrast, in the country with a clinical guideline for first-episode schizophrenia, the policy facilitated structured care through case notification, follow-up, and treatment. Programs in academic settings relied on researchers and trainees volunteering for core tasks like assessments and therapy, due to limited budgets.

> *“Depending on the resources that we had available, if we have someone, a psychologist, that would be a volunteer, then we could provide psychotherapy for [patients]. But so that’s why it’s hard for us to follow a specific model as we don’t have people really hired specifically for this.” [Interview 8]*

The implementation phase revealed operational challenges. For CHR, these were under-resourced health systems, coordination challenges (e.g., limited referral pathways), and contextual factors like substance use complicating diagnosis. For FEP, defining onset was difficult when individuals arrived after long periods of untreated psychosis or unreliable antipsychotic use records. Some participants, therefore, preferred broader terms like “untreated psychosis” or “early-onset psychosis.” Retention of service users was also a challenge, with many disengaging after initial symptom improvement. Programs often provided non-protocolized care based on resources and individual needs.

> *“We registered this participant as a patient here at the [institution], and we started treating the participants and there was no standard, it was more like on an individual basis.” [Interview 8]*

As clinical and research programs developed, some joined multicenter EIP studies, mainly contributing to participant recruitment, but gaining networks, research capacity, and funds. Some initiatives received industry support. One regional initiative was highlighted for unifying EIP efforts across LAC, setting regional research priorities and generating publications. Still, challenges included focusing solely on EIP, competing research priorities, and limited funding mechanisms.

> *“Research in Latin America exists, there are funds. There are places that obviously have a greater offering; people from Brazil with FAPESP have good support; in Chile, ANID works; for example, COLCIENCIAS in Colombia also works. The Mexicans also works since they have the CONACYT. The problem is that generally all these funds are for intra-country financing. So, there’s no way to harmonize projects together, and that’s where we fall.” [Interview 22]*

### Sustainability phase

EIP initiatives followed various sustainability trajectories revealing implementation challenges in LAC (Fig 5). The COVID-19 pandemic influenced both their development and long-term viability. A clinical program was discontinued due to administrative disruptions and service reorganization; another due to difficulties in identifying and retaining service users, both exacerbated by the pandemic.

**Fig 5.**
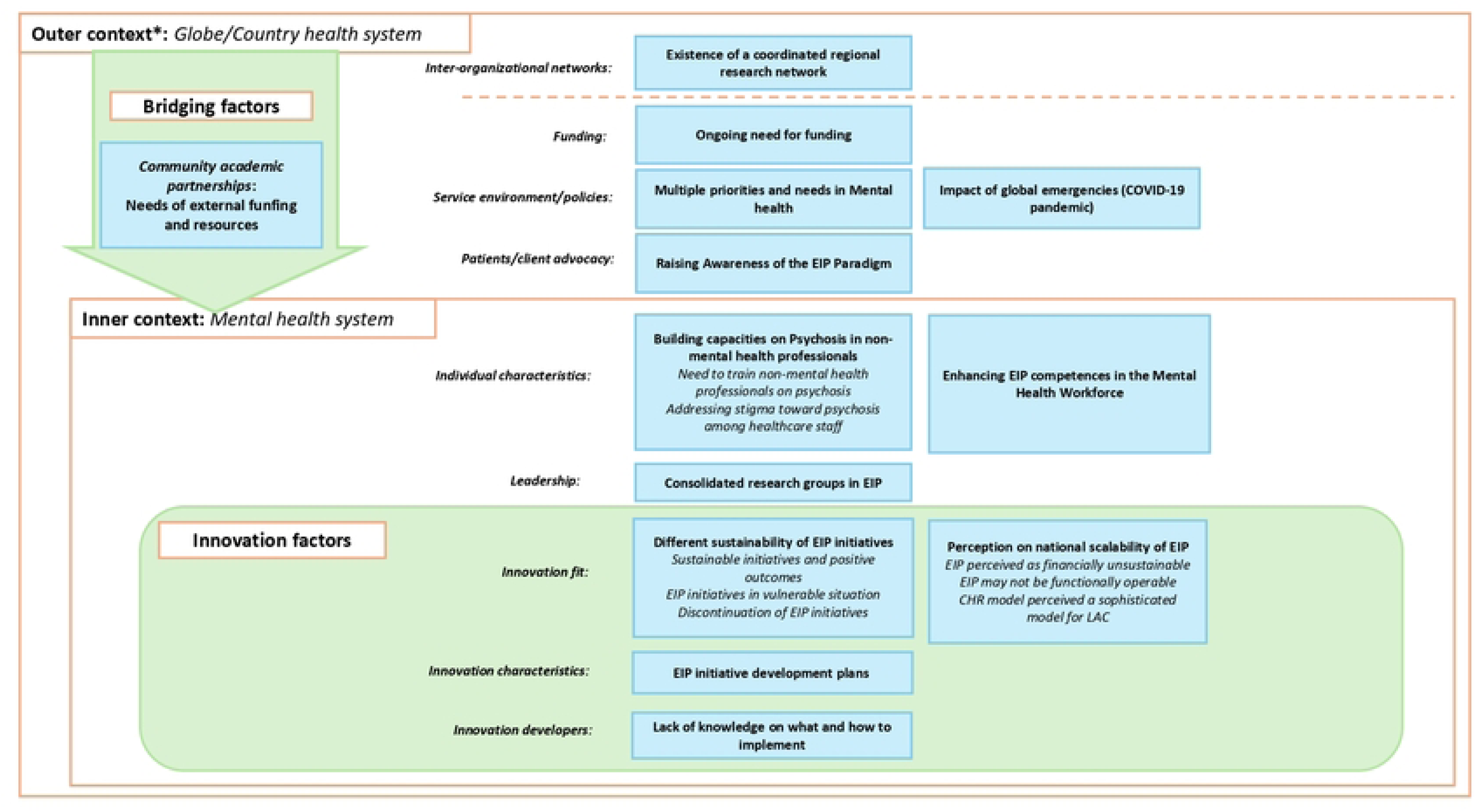
Themes and subthemes identified during the Sustainability phase of EIP initiatives (n = 26). *The outer context comprises two dimensions, separated by the dashed orange line: global context and national health system.

> *[EIP clinical program] around 2022, we closed it because we didn’t have so many volunteers. We didn’t have so many patients. And we decided then to focus on the first-episode program.” [Interview 8]*

At interview time, sustainable initiatives included two clinical guidelines, eight clinical programs, and four research programs. The guideline in the strong primary care–based country was seen as essential for enabling and sustaining related initiatives, whereas a similar guideline was deemed unfeasible in another country due to limited resources and staffing. This contrast underscores that policy, without proper resource allocation, has limited impact. Some initiatives remained fragile with unstable funding and staff shortages. Others had benefited from institutional support and recognition (e.g., one initiative had been in place for 25 years), and had expanded regionally or used research to inform national policies. Both vulnerable and consolidated initiatives continuously sought resources.

> *“Funding is already gone, and we are looking for more funding. And the idea would be to, let’s say, scale the screening process and make it a good cost benefit about screening process. This is one of the main goals, so that it turns sustainable.”[interview 6]*

Large EIP research projects in LAC depended on international funding. One persisted but faced uncertain system integration once funding ended; another’s continuation depended on new international funding; as local resources were insufficient for large-scale research. The one regional research initiative had depended on international funding, as national grants were typically limited to country-specific projects. It continued through strong regional collaboration despite the loss of funding.

> *“Although we currently do not have funding, we maintain a collaboration. That is, we always have collaborations in Latin America with contacts and knowing that we can also apply for things together. So, we are applying with different countries; now more have joined us… we are going to do something with Uruguayans and Argentineans, without money, less resources, but we are able to keep the network alive.” [Interview 22]*

Participants emphasized the need for sustained health sector action to support EIP initiatives, including reducing stigma; enhancing EIP training in academic programs; and improving mental health workers’ knowledge of psychosis and competencies to integrate care into non-specialized settings. As EIP remains novel, participants recommended raising visibility through media, conferences, and policy-/decision-maker engagement.

> *“Academia needs to be closer to those who are the public mental health organize the system. So I think academia needs to go outside the walls and try to influence mental health policies. Not only the training, because if we have the training, but we don’t have the service and we don’t have a protocol, but it’s adopted the whole country, we’ll do the same.” [Interview 8]*

Participants valued EIP but held concerns about its nationwide expansion, particularly of HIC-like stand-alone clinical services, given their perceived high costs and implementation challenges in contexts with widely unmet mental health needs and limited services. They also justified this given the lack of guidance on implementing EIP in complex LAC contexts.

> *“The difficult part is sometimes being able to implement it, right? I mean, I think that, in theory, we know that we have to treat it early and provide the best possible treatment. What is complicated, and perhaps not so clear to me, is how we are going to implement it across the country. But I would think that the will, at least theoretically, exists.” [Interview 13]*

### Alternative approaches to scaling EIP

As nationwide implementation of existing EIP programs was considered unfeasible in LAC, participants proposed context-responsive dissemination models. Some suggested reserving CHR models for research, given limited resources and the need to prioritize FEP care.

> *“Perhaps the best position of the CHR model for LMIC is to focus on research fundamentally, but it’s not possible in terms of wide implementation. It’s not so cost effective to implement this kind of model across a country because there are other conditions that require also attention.”[Interview 17]*

Strengthening existing primary care or youth mental health services was proposed as a platform for identifying, managing, or referring CHR cases, as these are already integrated into national systems and better positioned for early detection.

> *“I think it would be important to involve other institutions that work on mental health issues, which are not third level…that treat patients already with a diagnosis…for example, like a service that was implemented a few years ago, which is a hospital of emotions and treats young people, adolescents and young adults. So they offer psychological services and I think that working with them would be a very good option.” [Interview 1]*

Several participants recommended implementing FEP clinical programs in tertiary care settings, specialized institutions, or hospitals in major cities where services already exist, while also promoting early intervention in regional areas with limited services and trained staff.

> *“I think they can be done in all the major Latin American capitals, yes. I think they should and can be done. But also balancing it with the fact that in the regional cities there is a clear lack of psychiatrists, lack of development of services and awareness. So, it is not really feasible to be able to make an early intervention service that is more extensive, but probably rather to raise awareness of the issue and try to make an early recognition, a relatively benign intervention in more general services.” [Interview 22]*

Others proposed developing EPI protocols or care standards to guide service delivery across system levels, and staff training for implementation and sustainability.

> *“I would say that the two needs is to organize first episode protocol in the health system, [country name], using the existing network, and also to organize better the psychiatric emergence in the country.” [Interview 8]*

Some recommended innovative delivery strategies, like task-shifting and simplified care packages, reflecting concerns about the feasibility of resource-intensive HIC models in low-resource settings.

> *“We would have to think of a compact version of that without losing the principles. If we cannot include interventions to prevent suicide or cognitive remediation. We can include other types of cheaper, more flexible interventions, which can be task-shifting, which can be provided here and which people have experience.” [Interview 2]*

## Discussion

Our findings highlight the contextual realities, structural barriers, and adaptive strategies shaping EIP initiatives across different implementation phases and levels of the social ecology in LAC. The EIP paradigm in LAC has been translated into diverse initiatives, largely driven by individual motivation, modelled after foreign programs, and constrained by local resources. While participants valued EIP, they cautioned against stand-alone EIP programs in LAC due to limited resources, instead proposing contextually grounded, resource-sensitive alternatives to ensure feasibility and effective scaling across LAC.

### Toward a broader approach to EIP in LMICs

It is well established that replicating models developed in HICs is often unfeasible in LMICs [25,47]. Some have proposed implementing only “key ingredients” of these interventions [25]. However, this assumes clearly defined core components that work across contexts, an assumption insufficiently defined. This one-dimensional framing also overlooks the potential of alternative strategies in LAC and may partly explain the stagnation of EIP implementation in many LMICs. Our findings underscore the value of diverse initiatives, whether standardizing practices, prioritizing specific populations, or addressing varied needs, thus calling for a more flexible conceptualization of EIP to advance it.

Beyond implementing programs, LMICs must foster complementary structures to enhance psychosis care. Unlike in HICs, guidelines, technical standards, and research [16,48,49] are largely absent in most LMICs [47]. Consequently, there is often no robust legal, educational, or evidence-based foundation to support the implementation of EIP programs in these settings. These structures are essential to support broader engagement in and sustainability of EIP, by leveraging local strengths such as strong primary care systems, community networks, and advocacy groups.

### Context shaping EIP initiatives

Our results suggest that EIP programs in LAC have been shaped by the same structural conditions and resource limitations that define mental health care in LMICs: low policy and funding priority, scarce or fragile funding, and reliance on individual initiatives or external support. This is unlike HICs, like the U.K. Australia, Denmark, Singapore and Canada [11,50–53], where strong political and financial commitment enabling widespread implementation and sustainability of EIP.

The provision of psychosocial interventions in LMICs is recognized as highly challenging due to limited resources [54,55]. EIP services in most LAC countries faced similar barriers. Although policies often stated that psychosocial care should be available nationwide, participants acknowledged that broader implementation was unfeasible due to a shortage of trained human resources or because services were concentrated in tertiary care. Although no formal fidelity evaluations were reported, care was generally described as non-protocolized/non-specialized and only partially aligned with international recommendations and local aspirations and needs.

### International influences

Most EIP initiatives were conceptualized on foreign models, aided by guidelines, implementation manuals, and connections with implementers from HICs. This externally driven approach may have introduced a desire to emulate foreign practices and a missed opportunity to fundamentally incorporate contextual knowledge and culturally relevant practices. Systematic cultural and content adaptation was not formally pursued in any initiative. Only public policies showed some degree of co-design with service users. Instead, adaptations emerged pragmatically, based on implementers’ experience and resources, a process also reported in other LMICs [56].

Interestingly, in the cases with formal North–South collaboration and external funding, there was an emphasis on capacity building and cultural sensitivity. In one case, the international funder required that institutional leadership be established in the South. In another, the funding call requested the inclusion of culturally sensitive practices and local stakeholders. While such actions by funders should continue [57], they come with a risk of tokenistic practices [58]. Dependence on foreign funding may also discourage local investment; and external funds can be abruptly withdrawn due to shifting political or institutional priorities. Future efforts must therefore center LMIC agency and leadership in EIP [14].

### Perspectives on scaling EIP

This study has important implications for EIP implementation in LAC. In fragmented, under-resourced, urban-centered mental health systems, participants viewed scaling traditional EIP clinical program models as largely unfeasible and difficult to replicate from HICs. They called instead for flexible, context-specific strategies that integrate EIP into national agendas and align with existing capacities, while addressing structural inequities, strengthening the workforce, and promoting mental health literacy to reduce stigma and improve understanding of psychosis.

Participants’ proposals for dissemination of EIP were experienced-informed and appear feasible within low-resource environments, and could be structured to inform resource allocation, policy and workforce training. Strategies, such as task sharing and task shifting, have already been successful in scaling mental health interventions in LMICs [59,60]. Regardless of the initiatives implemented, psychosis care must be included in universal health coverage frameworks to ensure that the population has access to services and financial protection against associated costs. The experience of LMICs like China and Brazil suggests that this measure can improve outcomes and reduce care gaps [61]. Future EIP implementation must be centered more strongly on the voices of people with lived experience and families, which is currently missing despite such involvement being a rights-based imperative that can enhance uptake, innovation and advocacy [62].

### Limitations and strengths

This study focused on implementers’ perspectives. Future work should integrate the perspectives of services users and families. Second, the implementers were predominantly men, which may have shaped findings around gendered influences on needs and implementation pathways. Third, some initiatives may have been overlooked, particularly of countries with low research capacity, although a wide definition for EIP initiatives and multiple identification strategies were used. Finally, by examining only EIP actors, other approaches to address psychosis and mental illness in LAC may have been missed, some of which may be more locally grounded and more scalable on their own or as elements of EIP. Despite these limitations, the richness and consistency of data across countries strengthen credibility of our findings.

This is the first qualitative study to comprehensively examine EIP implementation across multiple countries in LAC, covering 26 initiatives across 10 countries, providing a comprehensive regional perspective. It also captured the evolving nature of EIP in the region. For example, a technical standard moved from draft to rollout during the study. Triangulation and member checking enhanced the trustworthiness of the findings.

Implementation science remains limited in EIP and more generally in LMICs. Guided by the EPIS framework, our study makes important methodological and substantive contributions. Still, it did not fully capture factors such as supernatural explanations of psychosis, poverty, and other social determinants. We suggest viewing implementation science frameworks as adaptable tools, echoing previous calls to add domains like resource constraints and system characteristics in LMIC implementation studies [56].

This study shows how the EIP paradigm has been translated into diverse, locally adapted initiatives across LAC, despite constraints, limited funding, and uneven political support. Some achieved progress and others struggled with sustainability. Findings underscore the need to move beyond replicating HIC models toward a broader range of initiatives aligned with local priorities and capacities. Implementers’ proposals—integrating EIP into youth mental health and primary care, promoting task-shifting, simplified care packages and early psychosis literacy—offer feasible strategies for scaling early psychosis care in resource-limited settings.

## Data Availability

The datasets generated and analyzed during the current study are not publicly available, as they could reveal the identity of participants. However, they are available from the corresponding author upon reasonable request.

## Acknowledgements

We would like to thank all participants in this study for their time and for showing genuine interest in the conduct of this work.

## Supporting information

**S1 Table. Standards for Reporting Qualitative Research**

